# COVID-19 Vaccine Failure in Chronic Lymphocytic Leukemia and Monoclonal B-Lymphocytosis; Humoral and Cellular Immunity

**DOI:** 10.1101/2021.10.28.21265549

**Authors:** Yandong Shen, Jane A. Freeman, Juliette Holland, Ann Solterbeck, Kartik Naidu, Asha Soosapilla, Paul Downe, Catherine Tang, Ian Kerridge, Lucinda Wallman, Nenna Van Bilsen, Vanessa Milogiannakis, Anouschka Akerman, Gabriela Martins Costa Gomes, Kerrie Sandgren, Anthony L Cunningham, Stuart Turville, Stephen P. Mulligan

## Abstract

Chronic lymphocytic leukemia (CLL) is associated with immunocompromise and high risk of severe COVID-19 disease and mortality. Monoclonal B-Lymphocytosis (MBL) patients also have immune impairment. We evaluated humoral and cellular immune responses in 181 patients with CLL (160) and MBL (21) to correlate failed seroconversion (<50AU/mL SARS-CoV-2 II IgG assay, antibody to spike protein, Abbott Diagnostics) following each of 2 vaccine doses with clinical and laboratory parameters. Following first and second doses, 79.2% then 45% of CLL, and 50% then 9.5% of MBL respectively remained seronegative, indicating 2 vaccine doses are crucial. There was significant association between post-dose 2 antibody level with pre-vaccination reduced IgM (*p*<0.0001), IgG2 (*p*<0.035), IgG3 (*p*<0.046), and CLL therapy within 12 months (*p*<0.001) in univariate analysis. By multivariate analysis, reduced IgM (*p*<0.0002) and active therapy (*p*<0.0002) retained significance. There was no significant correlation with age, gender, CLL duration, IgG, IgA or lymphocyte subsets. Anti-spike protein levels varied widely and were lower in CLL, than MBL, and both lower than normal donors. Neutralization activity showed anti-spike levels <1000AU/mL were usually negative for both an early viral clade and the contemporary Delta variant. There were 72.9% of CLL and 53.3% of MBL who failed to reach anti-spike levels >1000AU/mL. In a representative subset of 32 CLL patients, 80% had normal T-cell responses by IFNγ and IL-2 FluoroSpot assay. Failed seroconversion occurred in 36.6%% of treatment-naive patients, 52.9% treatment-naive with reduced IgM, 78.1% on therapy, and 85.7% on ibrutinib. Vaccination failure is very common in CLL, including early-stage disease.

**6 Key Novel Findings:**

1. **Comparison CLL vs MBL vs normal**
  - 45% of CLL and 9.5% of MBL fail to seroconvert with 2 doses of COVID-19 vaccine
2. **Neutralization assay**
  - SARS CoV-2 IgG levels <1000 AU/mL rarely associated with neutralization activity.
3. **COVID-19-specific T-cell function by FluoroSpot IFN-g and IL-2 production**
4. **IgG, A, M class and IgG subclass:** correlations by univariate and multivariate analysis
  - IgM (OR 7.29 p<0.0001), IgG2 and IgG3 subclass univariate significance
5. Correlation with **therapy** – ICT, targeted therapies, and those on Ig replacement
6. **High risk of vaccination failure for all CLL, including early-stage disease, and MBL**

**Key Points:** CLL and MBL show significantly impaired anti-spike antibody, viral neutralization, with cellular immune response to COVID-19 vaccination

Failure to seroconvert is associated with low IgM, IgG2, IgG3, and recent therapy; many CLL and MBL patients remain COVID-19 vulnerable

## Introduction

Chronic Lymphocytic Leukemia (CLL) is almost invariably associated with some degree of immune failure, that typically worsens over the course of the disease.^1-3^ About 85% of CLL patients ultimately develop hypogammaglobulinemia, but also a wide range of both humoral and cell-mediated immune defects.^2,3^ This predisposes to higher risk of infection^4,5^ and second malignancy.^6-8^ There is a higher frequency and severity of viral diseases such as herpes zoster,^9,10^ and for some, such as rhinovirus,^11,12^ there is impaired ability to clear the virus.

Severe Acute Respiratory Syndrome Corona-virus-type 2 (SARS-CoV-2) is a single-stranded RNA virus that causes human infection, and COVID-19 disease manifestations, range from asymptomatic through to severe respiratory failure and death.^13^ COVID-19 has a major impact in patients with CLL.^14,15^ Data from the European Research Initiative on CLL (ERIC) show in CLL patients with confirmed COVID-19, 79% developed severe disease and require hospitalization and 36.4% died.^16^ Even in recent waves of the disease, mortality rates of 11% have been typical.^17^ Prolonged live virus shedding of SARS-CoV-2 has also been documented in CLL.^18,19^

Since the advent of COVID-19 vaccines, CLL patients have been included in national and global vaccination programs. Early data indicate that failure of seroconversion and probable ongoing COVID-19 susceptibility is a problem in CLL.^20-22^ For many, SARS-CoV-2 infection is likely resulting in major immediate risk of morbidity and mortality. We sought to better understand the immune impairments that predict failure of seroconversion by analyzing B and T-cell immune responses to COVID-19 vaccines and correlating with a wide range of clinical, therapeutic, and biological characteristics including quantification of immunoglobulins (G, A, M, IgG subclasses), lymphocyte subsets, and COVID-19-specific neutralization antibody and T-cell responses.

## Methods

The study was approved by the Northern Sydney Local Health District Human Research Ethics Committee (approval number: LNR/14/HAWKE/181) and all patients provided an informed consent. The diagnosis of CLL and MBL were according to iwCLL guidelines.^23^ Vaccination occurred through the Australian Government vaccination program (www.health.gov.au) where the main initial vaccine availability was internationally and domestically produced Vaxzevria (AstraZeneca AZ), and more recently, Comirnaty (Pfizer) and Spikevax (Moderna) mRNA vaccines.^24^ Blood samples (FBC, biochemistry, immunoglobulins G, A, M, IgG subclasses, phenotyping and COVID-19 antibody levels) were taken pre-vaccination, then following vaccination doses 1 (D1) and 2 (D2) approximately 2 to 4 weeks following each dose. CLL therapy was avoided unless essential or given in the periods when there was no community COVID-19 transmission.^25^

IgG (6.5-16g/L), A (0.4-3.5g/L) and M (0.53-3g/L) were performed on the Siemens Atellica. IgG subclasses (IgG1, 3.82-9.29g/L; IgG2, 2.42-7g/L; IgG3, 0.22-1.76g/L; IgG4, 0.04-0.86g/L) were performed using Optilite IgG subclass kits (The Binding Site). Analysis of IgG and IgG subclass data excluded all patients on immunoglobin replacement therapy (IgRT). Flow cytometry was performed on a Navios 10 color instrument with Beckman-Coulter fluorochrome-labelled monoclonal antibodies.

SARS-CoV-2 IgG II Quant assay^®^ (Abbott Diagnostics) was performed in accordance with the manufacturer’s instructions. This chemiluminescent microparticle immunoassay detects IgG antibodies to the spike receptor-binding domain (RBD) of the S1 subunit of the spike protein of SARS-CoV-2. The SARS-CoV-2 antigen-coated paramagnetic microparticles bind to the IgG antibodies against the virus’ spike protein in human serum and plasma samples. The resulting chemiluminescence in relative light units (RLU) following the addition of acridinium labeled anti-human IgG (mouse, monoclonal) in comparison with the IgG II calibrator/standard indicates the strength of response, which reflects the quantity of RBD IgG present. Fifty arbitrary units per milliliter (50AU/mL) and above in this test are considered positive. For this study, we also examined the degree of anti-spike protein response with arbitrary strata of <50 (negative), 50-249 (weak), 250-999 (moderate), 1000-4999 (strong), 5000-9999 (high), and >10000 (very high).

### SARS-CoV-2 live virus neutralization assay

HEK-ACE2/TMPRSS cells (Clone 24)^26^ were seeded in 384-well plates at 5×10^3 cells per well in the presence of the live cell nuclear stain Hoechst-33342 dye (NucBlue, Invitrogen) at a concentration of 5%v/v. Two-fold dilutions of patient plasma samples were mixed with an equal volume of SARS-CoV-2 virus solution (1.25×10^4 TCID50/mL) and incubated at 37°C for 1 hour before adding 40μl in duplicate to the cells (final MOI = 0.05). Viral variants used included the variants of concern; Delta (B.1.617.2), as well as ‘wild-type’ control virus (B.1.319/D614G strain) from an early circuiting 2020 clade (B.1). Plates were incubated for 24 hours post infection and entire wells were imaged by high-content fluorescence microscopy, cell counts obtained with automated image analysis software, and the percentage of virus neutralization was calculated with the formula: %N = (D-(1-Q)) × 100/D, as previously described.^26^ An average %N>50% was defined as having neutralizing activity. In studies of convalescent patients, 96% of COVID-19 infected, convalescent patients reach a titer of 1/40 at their peak and is therefore used as the benchmark titer in this study.^26^ The mean titer of the WHO G serology standard in this assay for the above B1 clade virus is 1/637.

### IFNg/IL-2 Fluorospot Assay

PBMCs isolated from whole blood by Ficoll-Hypaque density gradient centrifugation were seeded in T-cell interferon gamma (IFNγ)/interleukin-2 (IL-2) dual color fluorospot plates (Mabtech). For patients with a CD3% within the normal reference interval, 250,000 cells per well were plated. For patients outside the reference interval, the cell number was normalized according to the patient’s CD3%, derived by flow cytometry on whole blood, to give either 100,000 (for low CD3%) or 220,000 (for high CD3%) T cells in a maximum of 400,000 cells per well. Where a patient’s CD3% was less than 20%, the maximum 400,000 cells per well was plated. Cells were incubated with an overlapping peptide pool spanning the complete S protein (2μg/mL; Miltenyi) for 18h at 37C. Negative control wells lacked peptides and PHA (10μg/mL; Vector Laboratories) was used as a positive control. Plates were developed according to the manufacturer’s instructions and read on a Mabtech Iris fluorospot reader. All tests were performed in duplicate, and the mean value was used for data representation.

### Statistical analyses

All analyses were conducted using SAS V9.4. The proportion of patients in various categories for response to COVID-19 vaccine and clinical variables are presented with exact 95% confidence limits. The association between COVID-19 vaccine response (negative/positive) after the second dose of vaccine and clinical variables was estimated using univariate logistic regression models, modelling the odds of a negative response, fitted with conditional exact methods to obtain exact P-values and 95% confidence limits. Exact methods were employed due to the small patient numbers in some categories. Multivariate logistic regression models, modelling the odds of a negative response after the second dose of vaccine, were fitted including multiple clinical variables found to be significant in univariate models. If two clinical variables were highly correlated (e.g. treatment within last 12 months and currently on treatment) only one variable was included. In addition, a final model was fitted including only terms statistically significant in the more inclusive multivariate model. Note, any analyses where pre-vaccination IgG levels (or IgG subclasses) were considered, patients currently treated with IgG replacement therapy were excluded.

## Results

### Patient characteristics

From 1^st^ March through to 22^nd^ October 2021, a total of 235 patients, 206 CLL and 29 MBL, were assessed. Statistical analysis only included patients with post D2 vaccine response available, i.e., 160 CLL and 21 MBL, in total 181 patients. The patient baseline demographic characteristics are shown in Table 1. The median age was 71.5 years for CLL, and 71 years for MBL. The proportion of males for CLL was 56.3%, and for MBL was 38.1%. The time from CLL diagnosis to vaccination ranged from <1 to 32 years with a median of 10 years.

**Table 1.** Patient characteristics.

| **Total number of patients (n=181)** | **CLL (n=160)** | **MBL (n=21)** |
| --- | --- | --- |
| Males | 56.3% (90) | 38.1% (8) |
| Median age (years) | 71.5 (22-94) | 71 (50-86) |
| Median age at diagnosis (years) | 60 (18-85) | 68 (48-84) |
| **Vaccine** |  |  |
| AstraZeneca | 136 (85.0%) | 13 (61.9%) |
| Pfizer | 19 (11.9%) | 4 (19.0%) |
| Moderna | 1 (0.6%) | 0 |
| Unknown | 4 (2.5%) | 4 (17.4%) |
| **Cytogenetics** |  |  |
| Del 13q | 79/122 (64.8%) | 3/5 (60.0%) |
| Del 11q | 14/122 (11.5%) | 0/5 |
| Del 17p | 9/122 (7.4%) | 0/5 |
| Trisomy 12 | 17/122 (13.9%) | 1/5 (20.0%) |
| Unknown | 38 | 16 |
| **IGVH** |  |  |
| Mutated | 24 | - |
| Unmutated | 6 | - |
| Unknown | 130 | 21 |
| **Treatment status at vaccination** |  |  |
| Treatment naïve | 82 (51.3%) | 21 (100%) |
| On-therapy | 32 (20.0%) | 0 |
| Off-therapy in remission (CR or PR) | 40 (25.0%) | 0 |
| Off-therapy in relapse | 6 (3.8%) | 0 |
| **Ig replacement therapy** |  |  |
| Currently on | 24 (15.0%) | 0 |
| Prior but not current | 9 (5.6%) | 0 |
| Never on Ig replacement therapy | 122 (76.3%) | 21 (100%) |
| Unknown | 5 (3.1%) | - |
| **Anti-CD20 inhibitor in the last 12 months** |  |  |
| Yes | 9 (5.6%) | 0 |
| No | 151 (94.4%) | 0 |

There were 82 patients with CLL who were treatment-naïve, 40 previously treated in complete or partial remission, 6 with progressive disease pending therapy, and 32 on active therapy. Of the later there were 21 on ibrutinib, 2 on FCR (fludarabine, cyclophosphamide, rituximab), 1 bendamustine plus rituximab, 4 venetoclax (1 venetoclax/ibrutinib), and 4 on other therapies. The majority (both CLL and MBL) 149 received AZ while 23 received Pfizer, and 1 received Moderna for both D1 & D2, while for 8 patients, the type of vaccination was unclear. No difference in anti-spike antibody efficacy was identified with the different vaccines. No patients had COVID-19 infection prior to vaccination. There were no thromboembolic events cerebral or otherwise with the AZ vaccine, and no episodes of pericarditis or myocarditis reported with Pfizer.

### Antibody responses

Antibody responses to COVID-19 spike protein (anti-S antibody) using the SARS-CoV-2 II IgG assay Abbott Diagnostics assay as qualitatively negative or positive (i.e. IgG level<50AU/mL or ≥50AU/mL) after D1 and D2 respectively are shown in Table 2. In CLL patients (n=160), 79.2% were negative after D1 and 45.0% remained negative after D2. For the 21 MBL patients, 40.0% and 9.5%, respectively were negative after each dose, compared to 0% and 0% of 25 normal controls (i.e. 100% of 25 normal controls seroconverted after D1) (Table 3 and Figure 1A). No patient or control had an anti-nucleocapsid antibody response prior to vaccination, consistent with absence of COVID-19 infection (data not shown).

**Figure 1.**
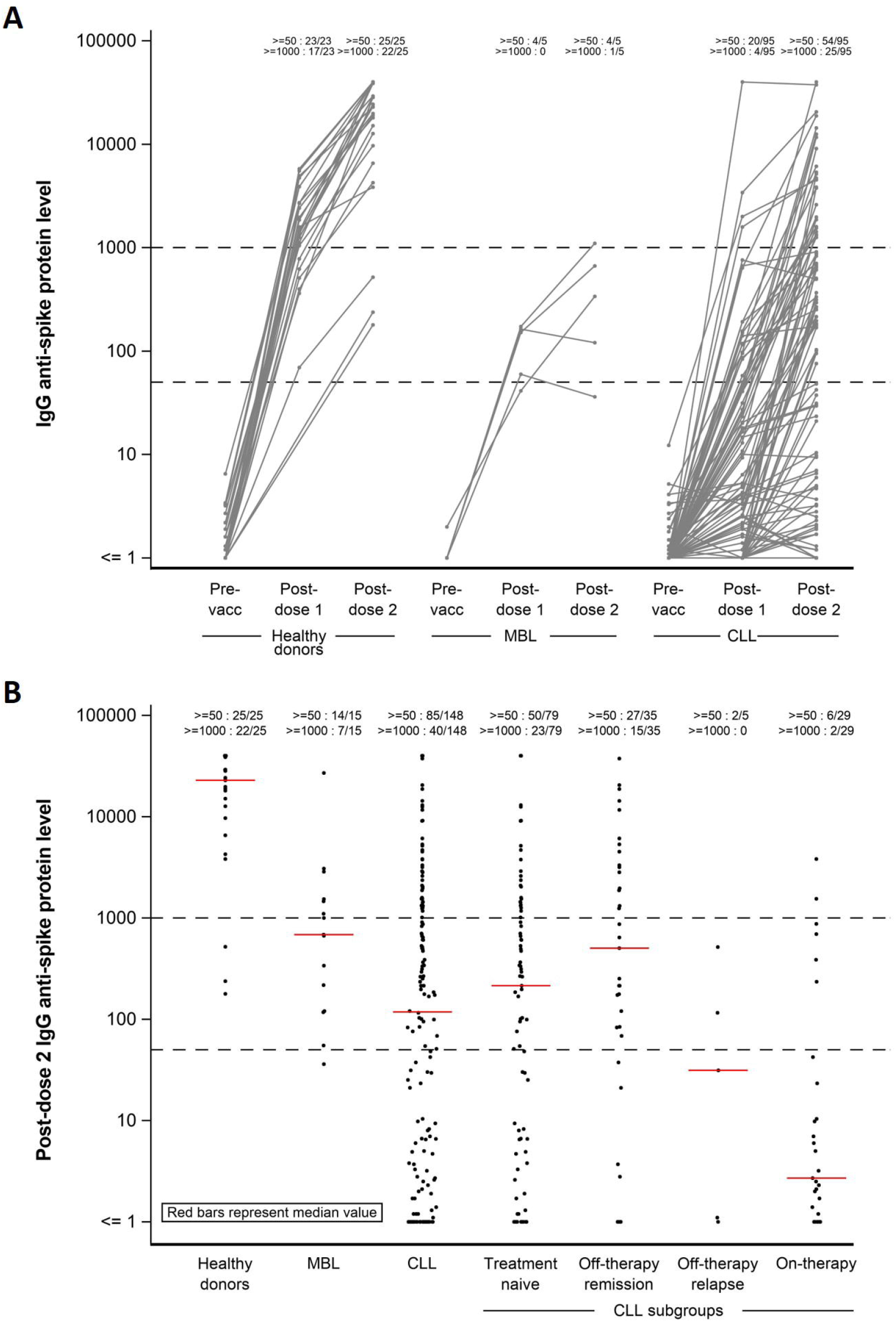
IgG anti-spike antibody levels post vaccination. (A) Changes in IgG anti-S levels pre-vaccination, post D1 and D2. Donors and patients with anti-S values <1 (including zero) is shown at <=1 on the graph. Values at the top of the graph at D1 and D2 display n/N, where n is the number of donors/patients with an anti-S value above the threshold, and N is the total number of donors/patients with a non-missing value. Thresholds are shown for >=50 and >=1000. (B) Post D2 IgG anti-spike antibody values. Red bars represent median titer value for each group.

**Table 2.** Anti-spike antibody response (qualitative: negative or positive [i.e. anti-S value >50]) rates in CLL and MBL.

| Anti-spike antibody response (negative or positive [i.e. anti-S value >50]) rates |  |  |  |  |
| --- | --- | --- | --- | --- |
| Population | Assessment | Response | Frequency/group total | Percentage (Exact CL) |
| All CLL and MBL | Post Dose 1 IgG spike protein response | Negative | 104/135 | 77.04% (69.02%, 83.83%) |
|  |  | Positive | 31/135 |  |
| CLL | Post Dose 1 IgG spike protein response | Negative | 99/125 | 79.20% (71.03%, 85.94%) |
|  |  | Positive | 26/125 |  |
| MBL | Post Dose 1 IgG spike protein response | Negative | 5/ 10 | 50.00% (18.71%, 81.29%) |
|  |  | Positive | 5/ 10 |  |
| All CLL and MBL | Post Dose 2 IgG spike protein response | Negative | 74/181 | 40.88% (33.65%, 48.42) |
|  |  | Positive | 107/181 |  |
| CLL | Post Dose 2 IgG spike protein response | Negative | 72/160 | 45.00% (37.14%, 53.05%) |
|  |  | Positive | 88/160 |  |
| MBL | Post Dose 2 IgG spike protein response | Negative | 2/ 21 | 9.52% (1.17%, 30.38%) |
|  |  | Positive | 19/ 21 |  |

**Table 3.** Anti-SARS-Cov2 Spike antibody level stratification in CLL, MBL patients and healthy donors post vaccination. Only patients with quantitative anti-spike antibody level available was included in this table.

|  | CLL |  | MBL |  | Healthy Donor |  |
| --- | --- | --- | --- | --- | --- | --- |
| Anti-spike antibody levels | Post D1 | Post D2 | Post D1 | Post D2 | Post D1 | Post D2 |
| <50 (negative) | 79.1% | 42.6% | 50.0% | 6.7% | 0.0% | 0.0% |
| 50-249 ('weak') | 14.8% | 14.2% | 50.0% | 26.7% | 4.3% | 8.0% |
| 250-999 ('moderate') | 2.6% | 16.2% | 0.0% | 20.0% | 21.7% | 4.0% |
| 1000-4999 ('strong') | 2.6% | 17.6% | 0.0% | 40.0% | 65.2% | 4.0% |
| 5000-9999 ('high') | 0.0% | 3.4% | 0.0% | 0.0% | 8.7% | 36.0% |
| >10000 ('very high') | 0.9% | 6.1% | 0.0% | 6.7% | 0.0% | 48.0% |

**Table 4.** Multivariate analysis for serologic response in CLL patients.

| **Population** | **Effect** | **Adjusted OR** | **95% CL** | **p-value** |
| --- | --- | --- | --- | --- |
| All CLL patients | **Pre-vaccination IgM**<br>Reduced vs Normal/high | 9.310 | 2.871-30.195 | 0.0002 |
|  | **On treatment vs Treatment naive/not on treatment** | 13.091 | 3.420-50.104 | 0.0002 |
| CLL no IgG treatment | **Pre-vaccination IgG2**<br>Reduced vs Normal/high | 1.190 | 0.512-2.767 | 0.6858 |
|  | **Pre-vaccination IgG3**<br>Reduced vs Normal/high | 2.105 | 0.802-5.530 | 0.1308 |

### Correlations with anti-S protein serologic response

Statistically significant associations were identified between failure to achieve an anti-S response with pre-vaccination low IgM (odds ratio [OR] 7.29, *p*<0.0001), low IgG2 (OR 2.52; p=0.035), low IgG3 (OR 2.68; *p*=0.046) and therapy within the prior 12 months (OR 5.20; *p*<0.0001) (Figure 2 and Supplementary Table 1). We did not find statistically significant associations with age, sex, or duration of CLL. Neither IgA (OR 1.69) or total IgG (OR 1.84; p=0.16) reached statistical significance. All analysis of IgG and IgG subclasses excluded patients on IgRT. There was no correlation with the CLL or MBL clonal population level, non-clonal B-cell numbers, total CD3+, nor CD4+ or CD8+ T-cells, and nor with memory CD4+ or memory CD8+ T-cell populations (data not shown). There was no correlation with CLL cytogenetics or immunoglobulin heavy chain variable region genes (IGHV) mutational status.

**Figure 2.**
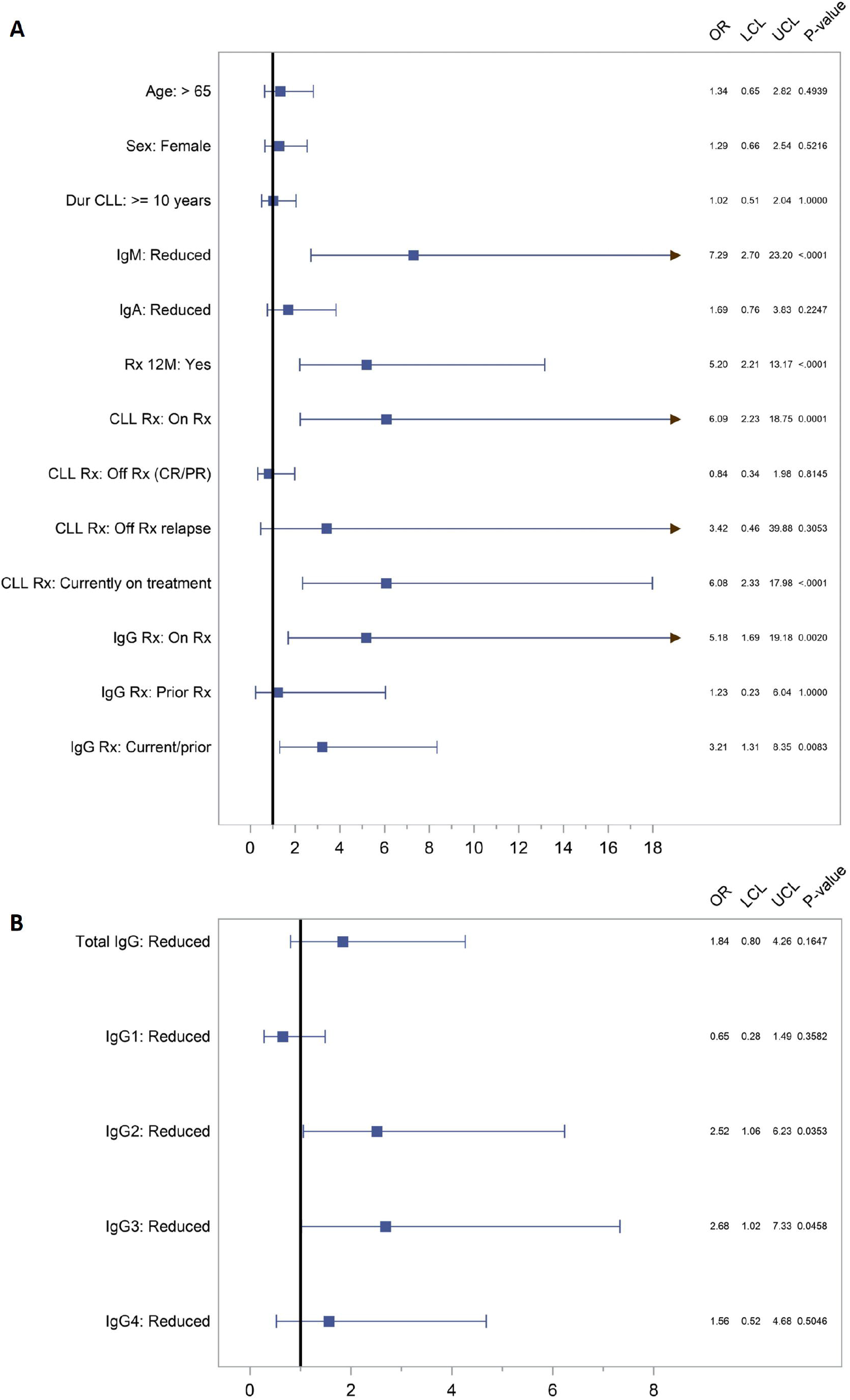
Odds ratio (OR) for no response to COVID-19 vaccine. (A) Anti-S responses after 2 vaccine doses by age, gender, duration of CLL, IgA, IgM, therapy within last 12 months, on-going therapy, off therapy in CR/PR, off therapy in relapse, and whether patient was on immunoglobulin replacement therapy, previously on, or both. (B) Anti-S responses after 2 vaccine doses by total IgG and IgG subclass, excluding patients on immunoglobulin replacement therapy. IgG anti-S response (negative or positive [i.e. anti-S value>50]).

Therapy in the prior 12 months was strongly associated with poor antibody response, with 74.4% failing to seroconvert (OR 5.20; p <0.0001) (Figure 2A and Supplementary Table 2). Some patients (n=6) with disease progression had their therapy deferred to minimize COVID-19 risk. For patients on active CLL-directed therapy, 78.1% remained seronegative after 2 vaccine doses (Supplementary Table 2). For those on active Btk-inhibition, mostly ibrutinib, 18 out of 21 failed to seroconvert (85.7%), and for those treated in the last 12 months, so did 7 of 9 receiving anti-CD20 (77.8%), 3 of 4 FCR, and 3 of 4 venetoclax. Interestingly, 2 of 2 patients on low dose prednisone (5mg daily) only for autoimmune cytopenia, and never on CLL-directed therapy had no seroconversion. Of CLL patients on IgRT for hypogammaglobulinemia with recurrent or severe infections, 77.3% (OR 5.18; *p*=0.002) failed to seroconvert (Figure 2A and Supplementary Table 2).

There were 82 (51.3%) treatment-naive patients 30 (36.6%) of whom failed to seroconvert. 52.9% of patients with reduced IgM as the only immunoglobulin class reduction failed to seroconvert.

### Multivariate analysis of serologic response in patients with CLL

In a multivariate analysis, we evaluated parameters associated with failure to seroconvert, namely pre-vaccination IgG2 and IgG3 levels (not on IgRT), IgM levels and on-going treatment. Pre-vaccination IgM levels (OR 9.31; *p*=0.0002) and on-going therapy (OR 13.09; *p*=0.0002) remained predictors of negative serologic response. Pre-vaccination IgG2 (OR 1.19; *p*=0.69) and IgG3 (OR 2.11; *p*=0.13) lost significance in multivariate analysis.

### Variation in anti-spike protein titers in CLL, MBL and normal

Anti-S levels in CLL demonstrated both a high proportion of negative results, and low levels compared to healthy controls (Figure 1). There was a wide range of anti-S levels from zero to >40,000. We divided these into six strata from negative to very high (Methods and Table 3) and measured neutralizing antibody in each stratum.

### Neutralization assay and anti-spike antibody levels

Neutralizing capacity of 30 patients (5 patients from each stratum) measured against both the original D614G and Delta SARS-CoV-2 variants are shown in Figure 3A and Supplementary Table 4. Of the 30 patients, neutralizing activity against the D614G strain was present in 12 (40%); 11 of those 12 had anti-S levels >1000, while 1 was 95AU/mL. Of the 30 patients, neutralizing activity against the Delta variant was seen in 9 (30%) and 8 had Anti-S levels >1000, and 1 was 875AU/mL. Hence, the majority of patients with neutralizing antibody had an anti-S level of >1000. Only 3 of 18 patients with anti-S level <1000 had detectable neutralizing activity.

**Figure 3.**
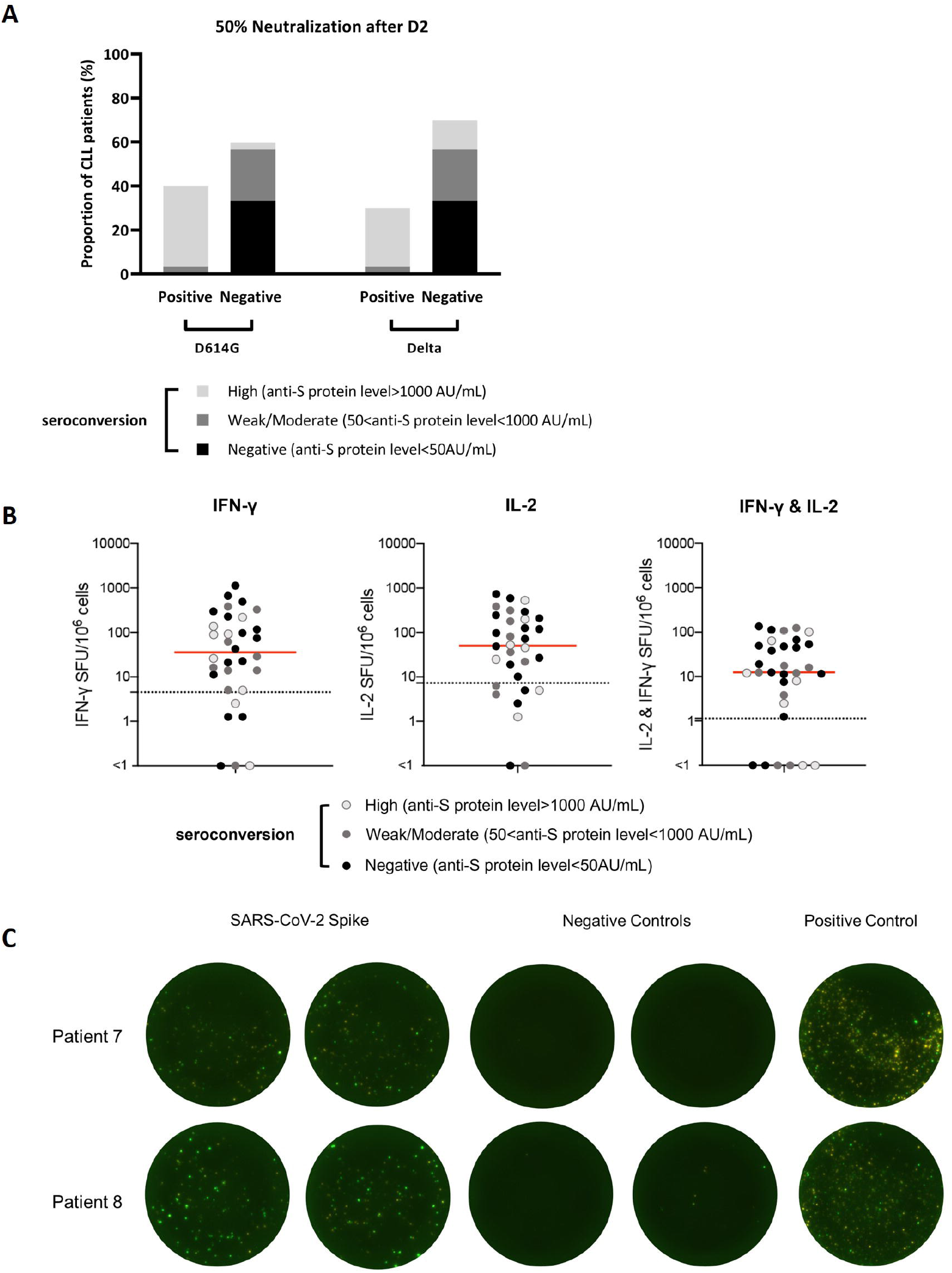
Post D2 vaccination neutralizing antibody and T-CLL function assays. (A) 50% neutralization against the D614G strain and the Delta variant of the SARS-CoV-2 after the second vaccination. Positive and negative results from neutralization assay were split into three stratifications of anti-S responses. (B) CLL patients mount functional T-cell responses to SARS-CoV-2 S protein after vaccination. Frequency of IFNγ and IL-2 producing T cells in response to an overlapping peptide pool covering the complete S protein in a dual color FluoroSpot assay. Each dot represents an individual participant. The red line denotes the median and the dotted line is the threshold for a positive response. SFU= spot forming units. n=32. (C) Image of FluoroSpot show duplicated results of patient 7 and 8. Green spots are IFNγ, and yellow spots are IL-2. Negative control lacked peptides and positive control contained Phytohemagglutinin (PHA).

In view of the almost invariable absence of neutralizing activity with anti-S level <1000AU/mL, we evaluated the parameters associated with failure to achieve an anti-S level of >1000. The data are shown in Supplementary Tables 1 and 3. After D2, 73.0% of CLL and 53.3% of MBL failed to reach an anti-S level >1000. Statistically significant associations between an anti-S level <1000, and IgM, IgG3 and treatment within the last 12 months remained, and IgA became statistically significant. The OR for total IgG was 2.75 (*p*=0.060), for IgM 2.81 (*p*=0.024), for IgA 3.30 (*p*=0.043), and for IgG3 3.84 (*p*=0.046). For patients on active therapy, 90% failed to achieve anti-S level >1000 (OR on therapy vs naïve/prior therapy 4.27, *p*=0.0659).

### Cell-mediated and COVID-19 specific T cell response

COVID-19 specific IFNγ and IL-2 responses were measured in 31 patients by the FluoroSpot assay across a range of anti-S antibody and clinical settings, many from the cohort with neutralizing assay results (Figure 3B). Overall, COVID-19 peptide stimulated T-cells from freshly collected blood samples showed essentially normal function in most patients. There was no correlation with either anti-S antibody production, nor with neutralizing activity. Of the 31 patients, 25 (80%) had a normal response, albeit of variable intensity, while 6 (20%) had a weak or negative response. Good T-cell function was present in several patients with prior and current therapy. There was no clear correlation between T-cell function and any clinical or therapeutic parameter.

## Discussion

CLL patients are virtually all immunocompromised to some extent^1,2^ and while SARS-CoV-2 vaccines have proven successful in protecting the general community, early data suggest that many patients with CLL fail vaccination^20-22^. In this study we examined patients across the spectrum of CLL, from those with a small peripheral blood clone, MBL^27,28^, to early stage CLL through to advanced stage and heavily treated CLL. Patients who fail to mount an antibody response likely remain COVID-19 susceptible and vulnerable to severe COVID-19 disease, hospitalization, intensive care unit (ICU) admission, and death.^15^ We therefore focused on this vulnerable group who failed to seroconvert examining both the humoral and cell-mediated immune response.

The study demonstrates a high proportion of CLL patients fail to achieve a positive anti-S level greater than the assay threshold of 50AU/mL, with 79.2% and 45% failing seroconversion after D1 and D2, respectively (Table 2). In MBL, 50% and 9.5% remained seronegative after D1 and D2. By contrast, a group of normal controls all achieved a positive anti-S level after D1, with much higher levels after D2. This emphasizes the importance of 2 vaccine doses in patients with CLL, and MBL. All patients with MBL by definition, and a high proportion of early stage CLL have few or no clinical problems with their disease, and many have minimal if any infection risk prior to COVID-19. Many of these patients do not mount a COVID-19 vaccine antibody response. In view of the indolent nature of their CLL, COVID-19 is now likely their highest risk of morbidity and mortality, far exceeding that from the CLL (or MBL) itself.

Parameters associated with failure to achieve a positive anti-S level in univariate analysis were pre-vaccination reduced IgM, IgG2, IgG3, and CLL therapy within 12 months (*p* <0.0001, 0.035, 0.046 and <0.001 respectively). There was no significant correlation with age, gender, CLL duration, IgG, IgA or lymphocyte subsets. Patients aged 65 years and older had a potentially meaningful higher risk of vaccine failure with an OR of 1.34, overall similar to data reported by Parry *et al*.^20^ and Herishanu *et al*.^21^.

Evaluating total IgG only in patients not on IgRT, the OR for pre-vaccination IgG was 1.84 but did not reach significance (*p*=0.17). There was a novel correlation with IgG2 and IgG3 and failure to seroconvert, but not IgG1, raising the interesting hypothesis that IgG2 and IgG3 may be more important than IgG1 for COVID-19 vaccine response.^29^ Furthermore, the lack of difference with response and IgG1 may contribute to why total IgG did not reach significance. Unlike 2 other studies^20,21^, we did not find significance with IgA, but this did emerge as significant when considering anti-S level >1000AU/mL as a strong serologic response.

Clinical variables significant in univariate analyses were considered in a multivariate model. Reduced IgM, prior treatment in the last 12 months, currently on treatment, current/prior IgG replacement therapy, reduced IgG2 and reduced IgG3 were all statistically significant at the p<0.05 level by univariate analysis. Due to the high correlation between any treatment in the last 12 months and currently on treatment, only “currently on treatment” was included in the model and enabled direct comparison with Israeli data^21^. Consistent with Herishanu *et al*.^21^, currently on treatment (OR 13.09; *p*=0.0002) and IgM (OR 9.31; *p*=0.0002), but not IgA, were predictors of serologic responses to vaccines. We did not find association between total IgG and vaccine responses. The OR for pre-vaccination IgG3 was 2.11 did not reach significance (p=0.13). Low IgG2 and low IgM were highly correlated as 45% of CLL patients had both (Supplementary data Table 5).

The strong association of failure to respond to vaccination with a low IgM level, often as a sole immunoglobulin class abnormality, is of concern as this is present in over a half of all CLL patients at diagnosis and in early-stage disease.^2^ This highlights the large numbers of CLL patients at ongoing risk, but also the importance of IgM and the primary immune response in developing antibodies.

Regarding CLL patients on therapy, as also demonstrated by others, there was a very strong association with vaccination failure in 74.4% (OR 5.20, p<0.001) of patients treated within the last 12 months, and 78% of those currently on active therapy failing to respond. Indeed, we found markedly impaired vaccine response with all forms of therapy with 18/21 ibrutinib,^30^ 7/9 CD20 antibody,^31^ 3/4 FCR and 3/4 venetoclax all failing to seroconvert.

Patients requiring IgRT as expected had very low vaccine response with 77.3% (OR 5.18; *p*=0.002) failing to develop antibodies. In Australia, the criteria for IgRT access for CLL patients are the presence of both hypogammaglobulinemia, and “recurrent or severe infection”. Hence this group is a surrogate for the “history of severe infection” group reported by Parry *et al*.^20^ and with a highly comparable response and failure rate.

Anti-S levels varied widely with many low, and of note lower in CLL, than in MBL, and in turn both were lower than healthy donors. Neutralization assays performed across the wide range of anti-S levels showed that neutralization activity was almost entirely restricted to those with an anti-S level >1000AU/mL. Using 1000AU/mL as the threshold of response, 73.0% of CLL and 53.3% of MBL failed to achieve this level. Also of note was that viral neutralization titers appeared weaker for the now globally dominant Delta variant compared to the original D614G strain. This is not unexpected as the vaccines were designed for D614G, but it is reassuring that significant activity against the Delta variant was demonstrated. T-cell responses appeared relatively intact, including those having received prior FCR and those on active therapy.^32^ Some patients appeared to have adequate T-cell numbers but failed to respond. Similarly, we found no significant correlation with total CD3+, nor CD4+ or CD8+ T-cells, and nor with memory CD4+ or memory CD8+ T-cell populations (data not shown).

Using an anti-S level >1000AU/mL as the threshold, parameters associated with negative response remain reduced pre-vaccination IgM, IgG3, and CLL therapy within 12 months (p<0.024, 0.046, and 0.041 respectively), but not for IgG2 (p=0.98). Interestingly and conversely, the association between reduced pre-vaccination IgA and failure to respond (OR 3.30, p=0.043) becomes statistically significant, and hence coincides with data reported by Parry *et al*.^20^ and Herishanu *et al*.^21^. Both those two reports employed the Roche Elecsys^®^ electrochemiluminescence immunoassay for measuring serologic responses while we used the Abbott Diagnostics assay. Differences in assay performance^33,34^ may contribute to discrepancies between our data and those of Parry *et al*.^20^ and Herishanu *et al*.^21^.

CLL and immunocompromised individuals are unable to clear certain viruses such as rhinovirus.^11,12^ For the much more serious COVID-19, long-term shedding of live virus has now been documented in CLL.^19,35^ The failure to develop anti-S antibodies or neutralizing activity is likely a major contributing factor to develop long-term viral replication and shedding.^35^ While the incidence of this issue remains unclear, its occurrence has major implications for CLL patients, their families, friends, and the broader community for infection control.

Some CLL patients do respond to vaccination as a recent second report from the Israeli group^36^ demonstrates, and this remains intact for at least 6 months. However, among CLL patients who fail to respond to 2 dose vaccinations, the response rates to third or more doses are likely to be low, and with low levels of anti-S,^37^ and this is consistent with our experience to date. Nevertheless, third and perhaps fourth vaccine doses are the easiest and most immediately available option and supported by our T-cell FluoroSpot data suggesting a functional T-cell anti-COVID-19 response in most patients. These T cell responses, especially CD8+ T cells, may be important in protection against severe lung disease, and T and B cell memory are important in durability of the vaccine response, probably especially in the ageing.^38,39^ Nevertheless, it is clear that there will be a significant group for whom no amount of vaccination will result in seroconversion.

For patients who fail to seroconvert, the ongoing management is likely to remain a challenge for some time and will almost certainly rely on passive immunity. In some settings, hyperimmune COVID-19 immunoglobulin^40^ is available but supply is likely to remain limited. Immunoglobulin replacement therapies may soon have anti-S antibodies detectable, however, the level of antibody and whether it confers any protective activity will be difficult to establish. Furthermore, the antibody levels will reflect vaccination rates in source plasma which is known to be low even in some settings with unrestricted vaccine access. Monoclonal antibody (MAb) preparations recently developed such as sotrovimab^41^ and the antibody combination (REGEN-COV casirivimab and imdevimab)^42^ are currently used in the setting of COVID-19 infection rather than prophylaxis due to the relatively short half-life (eg casirivimab and imdevimab, 32 and 27 days, respectively) of these MAb. The very recent data on the high-neutralization MAb cocktail, AZD7442 (co-administered tixagevimab and cilgavimab)^43^ engineered for a prolonged half-life of 9-12 months will make this and similar agents more practical as a prophylactic therapy for those patients who are unable to achieve protective antibodies by vaccination.

In conclusion, the rate of vaccination failure is very high in CLL, especially those on therapy, but also includes many with early-stage disease, and also individuals with MBL, i.e. patients for whom disease-related immune impairment previously had minimal clinical impact. Our data show there is a significant gap between seroconversion with anti-S antibody positivity and the presence of neutralizing antibodies, the latter of which are present in a much smaller proportion of MBL and CLL. It will be important to establish which elements of the immune system best predict Covid-19 protection as many appear to have an intact T-cell response. In any event, the standard precaution measures of masks, hand hygiene, social distancing, and vaccination of close family, friends and contacts will remain important and every CLL patient should remain conscious of these relatively simple precautions. The data from third and more vaccine doses will be important to evaluate as the information becomes available, as will the evaluation of MAbs and other potential passive immune strategies.

## Data Availability

All data produced in the present study are available upon reasonable request to the authors

## Acknowledgments

Robert Traficante, Statistical Revelations Pty Ltd, provided technical support in generating Figure 3 during preparation of this manuscript.

## Authorship Contributions

YS, JA, JH, KN, AS, PD, IK, LW, NB, SM collected the data. SM, JA, PD, IK confirmed the accuracy of the clinical data. VM, AA, GG, KS, AC, ST performed experiments and interpreted the data. YS and CT compiled the data for statistical analysis. AS performed the statistical analysis. YS and SM prepared the final manuscript, which all authors reviewed and approved.

## Tables and Figure legends

**Table 1.** Patient characteristics

**Table 2.** IgG anti-spike protein response (negative or positive [i.e. anti-S level>50]) rates in CLL and MBL.

**Table 3.** Anti-SARS-Cov2 Spike antibody IgG level stratification in CLL, MBL patients and healthy donors post vaccination. Only patients with quantitative anti-spike antibody level available was included in this table.

**Table 4.** Multivariate analysis for serologic response in CLL patients.

