## Supplemental tables/figure for "COVID-19 Vaccine Failure in Chronic Lymphocytic Leukemia and Monoclonal B-Lymphocytosis; Humoral and Cellular Immunity"

**Supplementary Table 1.** Anti-spike antibody response (negative or positive [i.e. anti-S value>1000]) rates in CLL and MBL.

| **Anti-spike antibody response (negative or positive [i.e. anti-S value >1000]) rates** | | | | |
| --- | --- | --- | --- | --- |
| **Population** | **Assessment** | **Response** | **Frequency/group total** | **Percentage (Exact CL)** |
| **All CLL and MBL** | Post Dose 1 IgG spike protein response | <1000 | 119/123 | 96.75% (91.88%, 99.11%) |
|  |  | >=1000 | 4/123 |  |
| **CLL** | Post Dose 1 IgG spike protein response | <1000 | 111/115 | 96.52% (91.33%, 99.04%) |
|  |  | >=1000 | 4/115 |  |
| **MBL** | Post Dose 1 IgG spike protein response | <1000 | 8/ 8 | 100.00% (63.06%, 100.00%) |
|  |  | >=1000 | 0/8 |  |
| **All CLL and MBL** | Post Dose 2 IgG spike protein response | <1000 | 116/163 | 71.17% (63.56%, 77.98%) |
|  |  | >=1000 | 47/163 |  |
| **CLL** | Post Dose 2 IgG spike protein response | <1000 | 108/148 | 72.97% (65.06%, 79.94%) |
|  |  | >=1000 | 40/148 |  |
| **MBL** | Post Dose 2 IgG spike protein response | <1000 | 8/ 15 | 53.33% (26.59%, 78.73%) |
|  |  | >=1000 | 7/ 15 |  |

**Supplementary table 2.** Anti-spike antibody responses after 2 vaccine doses by age, gender, IgG, IgA, IgM and IgG subclass (excluding patients on immunoglobulin replacement therapy for IgG and IgG subclass), and therapy within last 12 months. IgG anti-spike protein response (negative or positive [i.e. anti-S value>50]).

| **Variable** | **Category** | **Negative** | **Positive** | **Total** | **Conditional exact Odds ratio (95% confidence limits)** | **Exact P-value** |
| --- | --- | --- | --- | --- | --- | --- |
| **Age** | > 65 | 52 (47.3) | 58 (52.7) | 110 | 1.34 (0.65, 2.82) | 0.4939 |
|  | <= 65 | 20 (40.0) | 30 (60.0) | 50 |  | . |
| **Gender** | Female | 34 (48.6) | 36 (51.4) | 70 | 1.29 (0.66, 2.54) | 0.5216 |
|  | Male | 38 (42.2) | 52 (57.8) | 90 |  | . |
| **Duration of CLL** | < 10 years | 32 (45.1) | 39 (54.9) | 71 |  | . |
|  | >= 10 years | 35 (45.5) | 42 (54.5) | 77 | 1.02 (0.51, 2.04) | 1.0000 |
| **Prevaccination IgG*** | Reduced | 21 (50.0) | 21 (50.0) | 42 | 1.84 (0.80, 4.26) | 0.1647 |
|  | Normal/high | 27 (35.1) | 50 (64.9) | 77 |  | . |
| **Prevaccination IgM** | Reduced | 61 (58.1) | 44 (41.9) | 105 | 7.29 (2.70, 23.2) | <.0001 |
|  | Normal/high | 6 (15.8) | 32 (84.2) | 38 |  | . |
| **Prevaccination IgA** | Reduced | 22 (56.4) | 17 (43.6) | 39 | 1.69 (0.76, 3.83) | 0.2247 |
|  | Normal/high | 45 (43.3) | 59 (56.7) | 104 |  | . |
| **Prevaccination IgG1*** | Reduced | 18 (35.3) | 33 (64.7) | 51 | 0.65 (0.28, 1.49) | 0.3582 |
|  | Normal/high | 27 (45.8) | 32 (54.2) | 59 |  | . |
| **Prevaccination IgG2*** | Reduced | 32 (50.0) | 32 (50.0) | 64 | 2.52 (1.06, 6.23) | 0.0353 |
|  | Normal/high | 13 (28.3) | 33 (71.7) | 46 |  | . |
| **Prevaccination IgG3*** | Reduced | 16 (59.3) | 11 (40.7) | 27 | 2.68 (1.02, 7.33) | 0.0458 |
|  | Normal/high | 29 (34.9) | 54 (65.1) | 83 |  | . |
| **Prevaccination IgG4*** | Reduced | 10 (50.0) | 10 (50.0) | 20 | 1.56 (0.52, 4.68) | 0.5046 |
|  | Normal/high | 35 (38.9) | 55 (61.1) | 90 |  | . |
| **Any CLL treatment in last 12 months** | No | 43 (35.5) | 78 (64.5) | 121 |  | . |
|  | Yes | 29 (74.4) | 10 (25.6) | 39 | 5.20 (2.21, 13.2) | <.0001 |
| **CLL treatment status** | Treatment naïve | 30 (36.6) | 52 (63.4) | 82 |  | . |
|  | Off-therapy in remission (CR or PR) | 13 (32.5) | 27 (67.5) | 40 | 0.84 (0.34, 1.98) | 0.8145 |
|  | Off-therapy in relapse | 4 (66.7) | 2 (33.3) | 6 | 3.42 (0.46, 39.9) | 0.3053 |
|  | On-therapy | 25 (78.1) | 7 (21.9) | 32 | 6.09 (2.23, 18.8) | 0.0001 |
| **CLL treatment status** | Treatment naive/not on treatment | 47 (36.7) | 81 (63.3) | 128 |  | . |
|  | Currently on treatment | 25 (78.1) | 7 (21.9) | 32 | 6.08 (2.33, 18.0) | <.0001 |
| **IgG replacement therapy** | Not on treatment | 48 (39.3) | 74 (60.7) | 122 |  | . |
|  | Prior treatment not current | 4 (44.4) | 5 (55.6) | 9 | 1.23 (0.23, 6.04) | 1.0000 |
|  | On treatment | 17 (77.3) | 5 (22.7) | 22 | 5.18 (1.69, 19.2) | 0.0020 |
| **IgG replacement therapy** | Not on treatment | 48 (39.3) | 74 (60.7) | 122 |  | . |
|  | Current/prior | 21 (67.7) | 10 (32.3) | 31 | 3.21 (1.31, 8.35) | 0.0083 |

**Supplementary table 3**. Anti-spike antibody responses after 2 vaccine doses by age, gender, IgG, IgA, IgM and IgG subclass (excluding patients on immunoglobulin replacement therapy for IgG and IgG subclass), and therapy within last 12 months. IgG anti-spike protein response (negative or positive [i.e. anti-S value>1000]).

| **Variable** | **Category** | **Negative** | **Positive** | **Total** | **Conditional exact Odds ratio (95% confidence limits)** | **Exact P-value** |
| --- | --- | --- | --- | --- | --- | --- |
| **Age** | > 65 | 74 (74.7) | 25 (25.3) | 99 | 1.30 (0.56, 2.96) | 0.6160 |
|  | <= 65 | 34 (69.4) | 15 (30.6) | 49 |  | . |
| **Gender** | Female | 44 (72.1) | 17 (27.9) | 61 | 0.93 (0.42, 2.09) | 0.9919 |
|  | Male | 64 (73.6) | 23 (26.4) | 87 |  | . |
| **Duration of CLL** | < 10 years | 48 (72.7) | 18 (27.3) | 66 |  | . |
|  | >= 10 years | 57 (76.0) | 18 (24.0) | 75 | 1.19 (0.52, 2.72) | 0.8005 |
| **Prevaccination IgG*** | Reduced | 32 (84.2) | 6 (15.8) | 38 | 2.75 (0.96, 9.09) | 0.0602 |
|  | Normal/high | 50 (65.8) | 26 (34.2) | 76 |  | . |
| **Prevaccination IgM** | Reduced | 79 (80.6) | 19 (19.4) | 98 | 2.81 (1.13, 6.97) | 0.0242 |
|  | Normal/high | 22 (59.5) | 15 (40.5) | 37 |  | . |
| **Prevaccination IgA** | Reduced | 31 (88.6) | 4 (11.4) | 35 | 3.30 (1.03, 14.0) | 0.0428 |
|  | Normal/high | 70 (70.0) | 30 (30.0) | 100 |  | . |
| **Prevaccination IgG1*** | Reduced | 37 (74.0) | 13 (26.0) | 50 | 1.34 (0.53, 3.45) | 0.6325 |
|  | Normal/high | 38 (67.9) | 18 (32.1) | 56 |  | . |
| **Prevaccination IgG2*** | Reduced | 43 (71.7) | 17 (28.3) | 60 | 1.11 (0.43, 2.79) | 0.9806 |
|  | Normal/high | 32 (69.6) | 14 (30.4) | 46 |  | . |
| **Prevaccination IgG3*** | Reduced | 22 (88.0) | 3 (12.0) | 25 | 3.83 (1.02, 21.7) | 0.0464 |
|  | Normal/high | 53 (65.4) | 28 (34.6) | 81 |  | . |
| **Prevaccination IgG4*** | Reduced | 15 (78.9) | 4 (21.1) | 19 | 1.68 (0.47, 7.61) | 0.5691 |
|  | Normal/high | 60 (69.0) | 27 (31.0) | 87 |  | . |
| **Any CLL treatment in last 12 months** | No | 79 (68.7) | 36 (31.3) | 115 |  | . |
|  | Yes | 29 (87.9) | 4 (12.1) | 33 | 3.28 (1.04, 13.8) | 0.0410 |
| **CLL treatment status** | Treatment naïve | 56 (70.9) | 23 (29.1) | 79 |  | . |
|  | Off-therapy in remission (CR or PR) | 20 (57.1) | 15 (42.9) | 35 | 0.55 (0.22, 1.37) | 0.2236 |
|  | Off-therapy in relapse | 5 (100) | 0 (0.0) | 5 | 2.67 (0.47, I) | 0.1927 |
|  | On-therapy | 27 (93.1) | 2 (6.9) | 29 | 5.48 (1.20, 51.3) | 0.0215 |
| **CLL treatment status** | Treatment naive/not on treatment | 81 (68.1) | 38 (31.9) | 119 |  | . |
|  | Currently on treatment | 27 (93.1) | 2 (6.9) | 29 | 6.28 (1.45, 57.2) | 0.0073 |
| **IgG replacement therapy** | Not on treatment | 82 (70.7) | 34 (29.3) | 116 |  | . |
|  | Prior treatment not current | 7 (77.8) | 2 (22.2) | 9 | 1.45 (0.26, 15.0) | 0.9835 |
|  | On treatment | 16 (84.2) | 3 (15.8) | 19 | 2.20 (0.57, 12.5) | 0.3454 |
| **IgG replacement therapy** | Not on treatment | 82 (70.7) | 34 (29.3) | 116 |  | . |
|  | Current/prior | 23 (82.1) | 5 (17.9) | 28 | 1.90 (0.63, 6.93) | 0.3239 |

**Supplementary Table 4.** 50% neutralization against the D614G strain and the Delta variant of SARS-CoV-2 with corresponding anti-spike protein levels

| **Gender** | **Anti-S protein IgG** | **Titer** | **Vaccine** | **Neutralizing Ab titer against D614G** | **Neutralizing Ab titer against Delta** |
| --- | --- | --- | --- | --- | --- |
| M | Neg | 0 | AZ | <40 | <40 |
| M | Neg | 1.2 | AZ | <40 | <40 |
| M | Neg | 1.7 | Pfizer | <40 | <40 |
| F | Neg | 2.1 | AZ | <40 | <40 |
| F | Neg | 5 | AZ | <40 | <40 |
| M | Neg | 9.8 | AZ | <40 | <40 |
| M | Neg | 10.4 | AZ | <40 | <40 |
| F | Neg | 23.4 | AZ | <40 | <40 |
| F | Neg | 30.2 | AZ | <40 | <40 |
| F | Neg | 37.4 | AZ | <40 | <40 |
| M | Pos | 84 | AZ | <40 | <40 |
| M | Pos | 95.2 | AZ | 40 | <40 |
| M | Pos | 141.9 | AZ | <40 | <40 |
| M | Pos | 177.6 | AZ | <40 | <40 |
| F | Pos | 214.7 | AZ | <40 | <40 |
| M | Pos | 664.8 | AZ | <40 | <40 |
| F | Pos | 703.2 | AZ | <40 | <40 |
| F | Pos | 875 | AZ | <40 | 40 |
| M | Pos | 1340.5 | AZ | 40 | <40 |
| M | Pos | 1870.5 | AZ | <40 | <40 |
| M | Pos | 3771.1 | AZ | 160 | 80 |
| M | Pos | 4515.6 | AZ | 160 | 80 |
| M | Pos | 5159.5 | AZ | 80 | <40 |
| M | Pos | 5350 | AZ | 160 | 80 |
| M | Pos | 9081.4 | AZ | 80 | <40 |
| M | Pos | 14336.3 | AZ | 160 | 160 |
| M | Pos | 18781.3 | AZ | 320 | 160 |
| M | Pos | 20548 | AZ | 320 | 160 |
| F | Pos | 37586.4 | Pfizer | 320 | 160 |
| M | Pos | 40000 | AZ | 320 | 320 |

**Supplementary Table 5.** Correlation between pre-vaccination IgM and IgG2.

| **Pre-vaccination IgG2 (reduced vs normal)** | **Pre-vaccination IgM (reduced vs normal)** | | |
| --- | --- | --- | --- |
| **Frequency Row percentage Column percentage** | **Reduced** | **Normal/high** | **Total** |
| **Reduced** | 60 84.51 61.22 | 11 15.49 32.35 | 71 |
| **Normal/high** | 38 62.30 38.78 | 23 37.70 67.65 | 61 |
| **Total** | 98 | 34 | 132 |

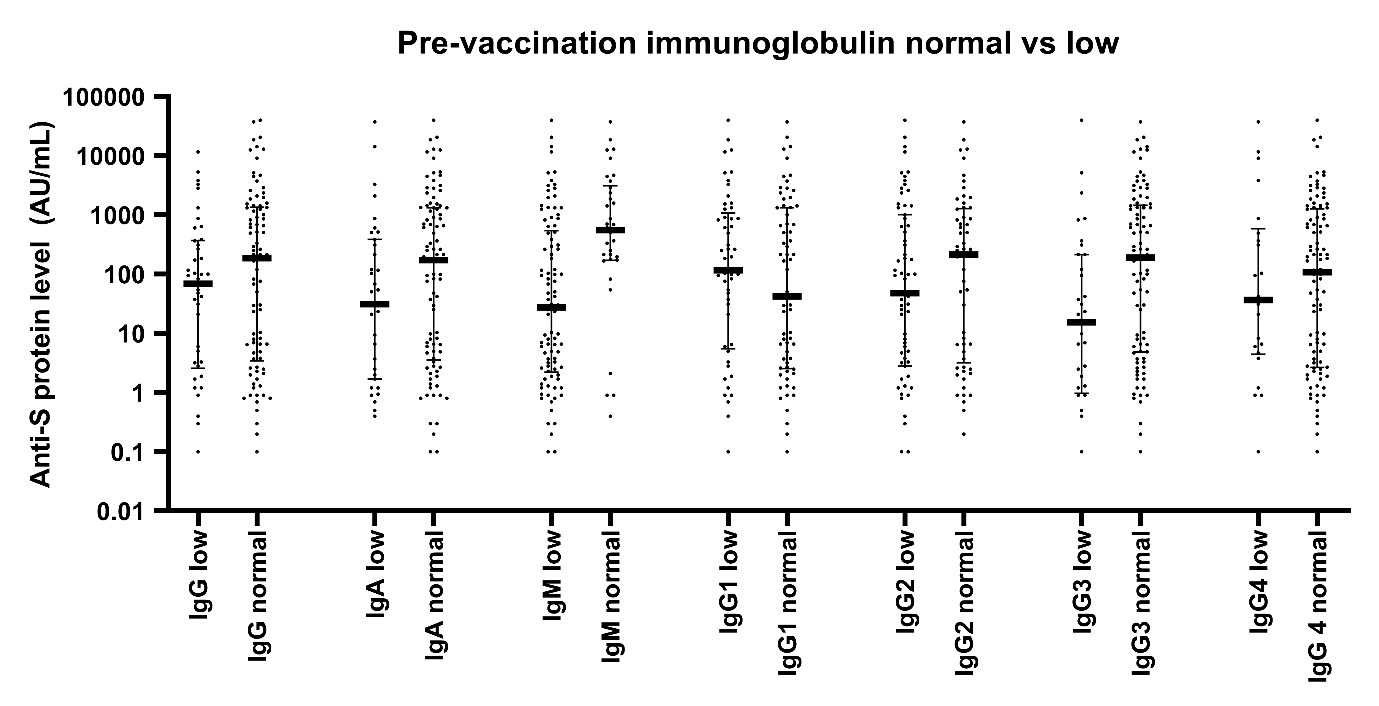

**Supplementary Figure 1.** Differences between post second dose vaccination anti-SARS-CoV2 S-protein levels in patients with normal and low pre-vaccination immunoglobulin levels.
